# Financial Health of Private Equity-Backed Ophthalmology and Optometry Groups: an Analysis of Debt Valuations

**DOI:** 10.1101/2022.11.12.22281833

**Authors:** Sarishka Desai, Rohail Memon, Evan Chen, Sachi Patil, Daniel Vail, Sailesh Konda, Ravi Parikh

## Abstract

**Purpose:** To identify debt valuations of ophthalmology and optometry private equity-backed group (OPEG) practices, which are a proxy for financial performance.

**Design:** Retrospective cohort study using the 2021 Business Development Company (BDC) Report and BDC quarterly/annual filings.

**Participants:** BDCs holding one or more ophthalmology/optometry group debt instruments.

**Methods:** The 2021 BDC Report was used to identify all BDCs actively filing annual reports (Form 10-Ks) and quarterly reports (Form 10-Qs) in 2021. The public filings of BDCs lending to OPEGs were searched from inception of an OPEG’s debt instrument in a BDC’s portfolio, and the amortized cost and fair value of each debt instrument were tabulated. A panel linear regression was used to evaluate temporal changes in debt valuations.

**Main Outcome Measures:** Trends in total debt held by OPEGs, trends in valuation (premium or discount) of OPEGs.

**Results:** A total of 2997 practice locations affiliated with 14 unique OPEGs and 17 BDCs were identified in 2021. Debt valuations of OPEGs decreased 0.46% per quarter over the study period (95% CI: −0.88 to −0.03, P = 0.036). In the COVID-19 pre-vaccine period (March 2020 – December 2020), there was an excess (additional) 4.93% decrease in debt valuations (95% CI: − 8.63 to −1.24, P = 0.010) when compared to prepandemic debt valuations (March 2017 – December 2019). Effects of COVID-19 on valuations stabilized during the pandemic post-vaccine period (February 2021 – March 2022), with no change in excess debt valuation compared to pre-pandemic baseline (0.60, 95% CI: −4.59 to 5.78, P = 0.822). There was an increase in practices that reported average discounted debt valuations from 20 practices (1.6%) associated with 1 OPEG to 1213 practices (40.5%) (including 100% of newly acquired practices) associated with 9 OPEGs, despite stabilization of COVID-19 related excess (additional) debt.

**Conclusions:** Valuations of OPEG debt have declined significantly post-PE investment from March 2017 to March 2022. An excess (additional) decline in valuations was observed during the COVID pre-vaccine period, with trends in excess debt valuations returning to baseline pre-pandemic levels by December 2021. Declining and discounted valuations of debt raise concerns about the financial viability of many PE-backed practices.

## Introduction

Private equity (PE) firms are asset managers that raise capital to obtain significant ownership in private companies. PE firms seek to improve the valuations of companies in their portfolios with the ultimate goal of selling these companies at a profit and, hopefully, generating above-market returns. Investments often have a short life span, from three to seven years from acquisition to turnover.^1^ PE acquisition continues to play a greater role in healthcare in the United States, particularly affecting procedure-based, outpatient-heavy fields such as dermatology, urology and ophthalmology.^2-4^

PE investment has implications not only on practice management strategy, but also the long-term economic wellbeing of practices. PE buyouts often involve raising substantial amounts of debt, often 70 to 80 percent of the purchase price, to meet the cost of acquisition.^5^ Debt financing allows PE firms to commit very little of their own capital (∼2% of purchase price) and achieve high internal rates of return (∼20%) in order to resell the company at a profit.^1^ However, this debt is carried by the practice, which shoulders the burden of repayment post-acquisition. The financial strength of a practice, along with market forces and interest rates, dictate its debt valuation.^6^ Interest rates have returned to pre-pandemic levels as of June 2021, minimizing the possibility external market factors are contributing to the valuation of debt.^7, 8^ Thus, the debt valuation of an ophthalmology/optometry PE-backed group (OPEG) can be used as a proxy for overall financial performance.^9-11^

Unfortunately, there is minimal reliable information about PE funds, the deals in which they engage, and the debt they hold. Business development corporations (BDCs), funds that invest in developing companies, are an exception.^12^ Unlike traditional PE funds which are limited to a select group of private investors, BDCs are traded on public stock exchanges and accessible to non-professional investors.^12^ Similar to publicly-traded companies, a BDC must file quarterly and annual reports with the Securities and Exchange Commission (SEC).^13^ Significant parameters related to debt, including the amortized cost of debt (net debt accumulated) and the fair value (estimated market value), are characterized in these reports.^14^ Studying debt reported by BDCs, as previously reported in the dermatology literature, can give us insight into trends of business acquisitions and correlated financial performance of OPEGs.^9^ The purpose of this study is to investigate how the debt valuations of OPEGs have fluctuated, if at all, since 2012, when our group previously reported a rapid increase in PE-backed acquisitions of ophthalmology and optometry practices^15^ Further, we sought to investigate how these debt valuations were impacted during the COVID-19 pandemic and COVID post-vaccine period, which demonstrated a normalization of ophthalmologic procedural volume to pre-pandemic levels.^16^

## Methods

### Data Collection

The BDC Report for 2021 was searched for publicly available BDCs, which were reviewed to find BDCs actively filing Form 10-Ks (annual business disclosure reports) and 10-Qs (quarterly reports).^14^ 79 BDCs with active quarterly and annual filings were identified, and the respective 10-Ks and 10-Qs were searched for the terms “ophthalmology,” “optometry,” “healthcare”, “eye, and “vision” to identify ophthalmology/optometry groups. Manual search of news sites, OPEG websites, and the financial database, Pitchbook, was conducted to identify practices backed by PE firms, and these groups were selected for inclusion in further analysis. Of note, Site for Sore Eyes and Sterling Optical are large, consolidated groups excluded from analysis as they are subsidiaries of Emerging Vision and not PE-owned.

The amortized cost and fair value of each OPEG’s debt instrument in a BDC’s portfolio was tabulated from the inception of the OPEG’s debt instrument in a BDC’s portfolio. Number of practice locations associated with each OPEG were identified by searching each platform’s website. To ensure consistency, locations of each OPEG as of July 2022 were reported.

### Valuation of OPEG Debt

The valuation of the total debt of an OPEG (premium or discount) at a given time was calculated by dividing the difference between the fair value and amortized cost by the amortized cost. Factors influencing the market value of debt include: interest rates, company performance/cash flow, value of company assets, and debt covenants.^9, 17^ A premium on the debt, meaning the fair value of the debt exceeds the amortized debt, means the debt is high quality and will continue to generate income until it matures.^18^ Debt valued at a discount, where the fair value is lower than the amortized cost, is a sign that the debt is of lower quality, and that the OPEG is at higher risk of defaulting on its debt.^19^

A panel linear regression was conducted to test for differences in debt valuations across fiscal quarters; individual OPEG debt obligations were used as the unit for the panel. Dummy variables representing the pre-pandemic period (March 2017 – December 2020), pandemic pre-vaccine period (February 2020 – December 2020) and pandemic post-vaccine period (February 2021 – March 2022) were included in the model to account for the specific impacts of the economic environment on debt valuations (excess debt due COVID-19 during those time periods). P < 0.05 was used as the cutoff for significance for all statistical tests. Statistical analysis was performed in June 2022 using Microsoft Excel 2016 and R, version 4.0.5 (R Group for Statistical Computing).

## Results

A total of 2,997 practice locations accounted represented by 17 BDCs and affiliated with 14 unique OPEGs were identified in 2021, with associated debt data available from March 2017 through March 2022. As more than one BDC can hold debt in the same OPEG, a total of 32 unique debt obligations were analyzed in this study. In February/March of 2022, the last quarter of data publicly reported during data analysis, a total of 492.9 million USD of amortized debt was held by the study sample. Total amortized cost of loans associated with an individual OPEG during the study period ranged from a low of 1.6 million USD (EyeSouth, February/March 2022) to 144.2 million USD (American Vision Partners, August/September 2021) (Figure 1).

**Figure 1.**
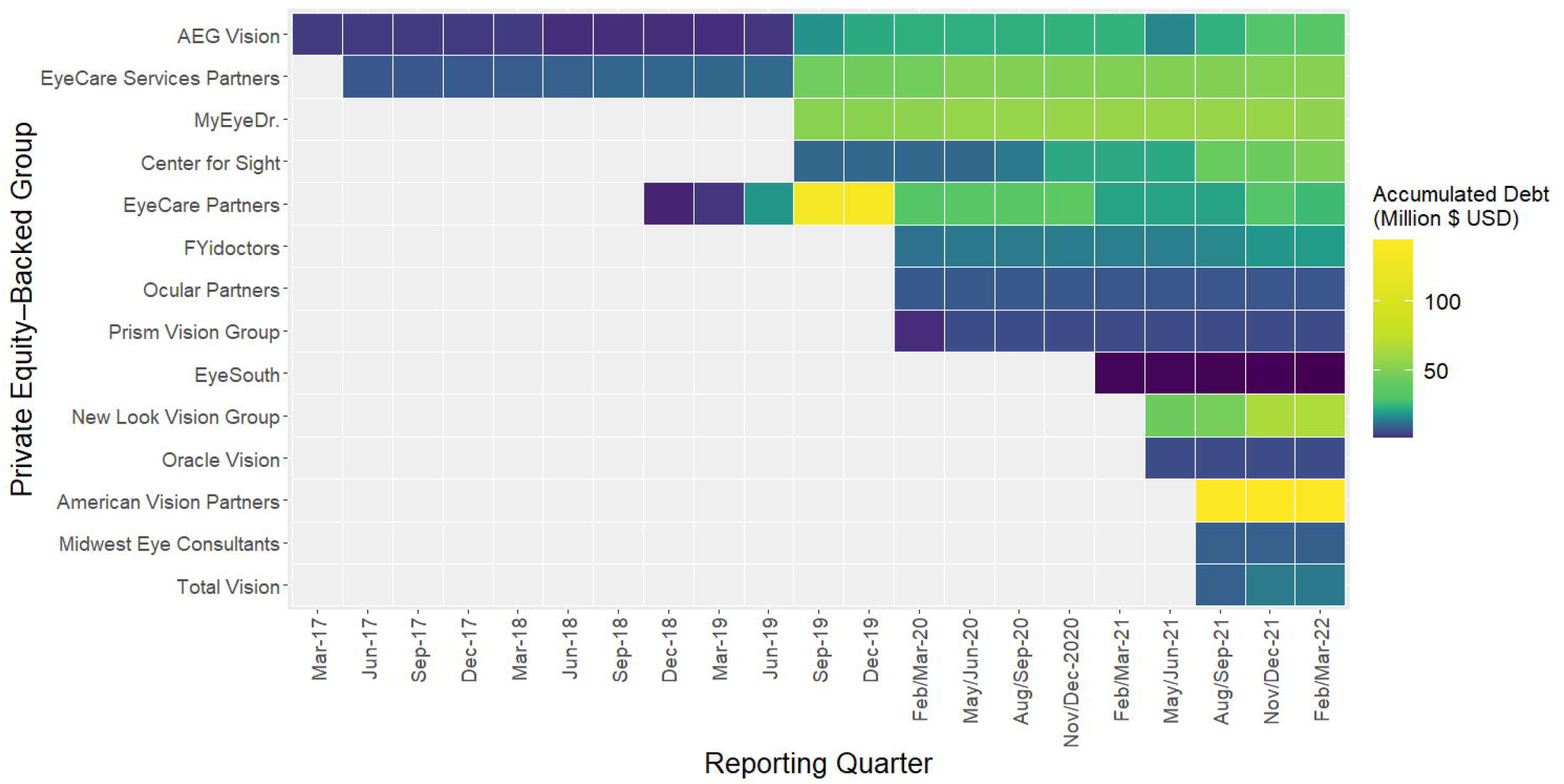
Total Amortized Cost of PE-Backed Ophthalmology and Optometry Debt Instruments

Of all reported debt in 2021, 8 debt instruments were acquired during the COVID pre-vaccine period (March 2020 to December 2020) and 12 additional debt instruments were acquired during the COVID post-vaccination period (February 2021 to March 2022). These acquisitions represent a 167% increase from the 12 OPEG debt instruments reported pre-pandemic, representing rising PE interest in ophthalmology and optometry practices. A plurality of BDCs, 47% (8/17), held only 1 debt instrument in their portfolio with 35% (6/17) holding 2 instruments, and 18% (3/17) holding 3 or more OPEGs in their portfolio during the study period. OPEGs were associated with over 2,997 practice locations throughout the United States and Canada as of July 2022 (Table 1).

**Table 1.**
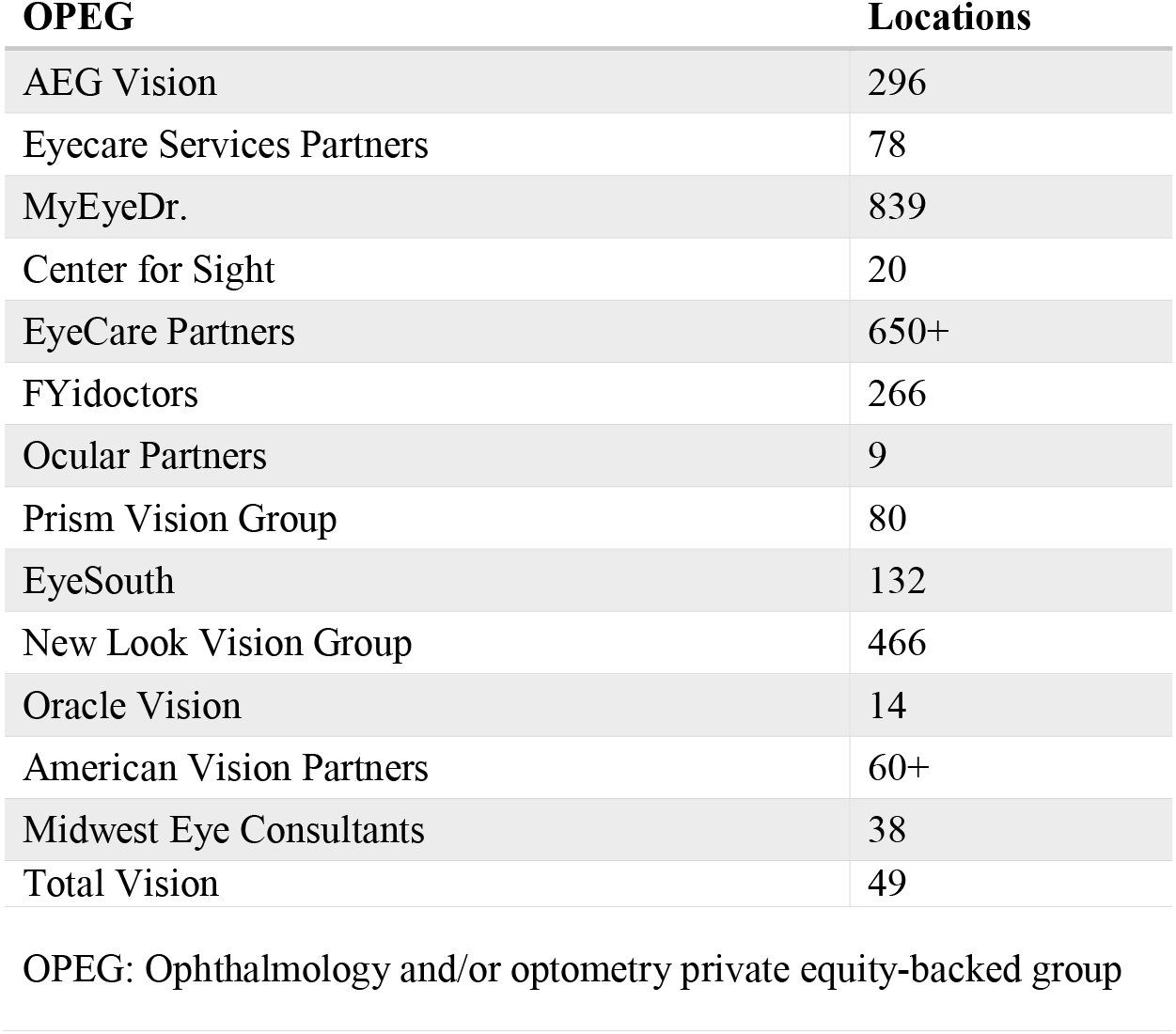
Number of OPEG Practice Locations (2022)

| **OPEG** | **Locations** |
| --- | --- |
| AEG Vision | 296 |
| Eyecare Services Partners | 78 |
| MyEyeDr. | 839 |
| Center for Sight | 20 |
| EyeCare Partners | 650+ |
| FYidoctors | 266 |
| Ocular Partners | 9 |
| Prism Vision Group | 80 |
| EyeSouth | 132 |
| New Look Vision Group | 466 |
| Oracle Vision | 14 |
| American Vision Partners | 60+ |
| Midwest Eye Consultants | 38 |
| Total Vision | 49 |

OPEG: Ophthalmology and/or optometry private equity-backed group

The valuation of debt instruments was found to be highly variable for many OPEGs during the study period (Figure 2). Panel linear regression analysis revealed the overall valuation of OPEG debt decreased 0.46% per quarter (95% CI: −0.88 to −0.03, P = 0.036) (Table 2). Despite this downward trend, OPEGs successfully maintained premium or close to par debt valuations from March 2017-December 2022 (pre-pandemic period), meaning practices were performing well financially. Acuity Eyecare (296 locations), MyEyeDr. (839 locations), Eyecare Services Partners (78 locations), and CFS Management (20 locations) all reported average pre-pandemic debt valuations between −0.1% and 1.3%.

**Table 2.** Panel Regression Model on OPEG Debt Valuation.

| **Predictor** | **Change in Valuation (%)** | **95% CI** | **P Value** |
| --- | --- | --- | --- |
| Time (Quarters) | -0.46 | -0.89 to -0.03 | 0.036 |
| Pandemic pre-vaccine<br>(March 2020 –<br>December 2020) | -4.93 | -8.63 to -1.24 | 0.010 |
| Pandemic post-vaccine<br>(February/March 2021 –<br>February/March 2022) | 0.60 | -4.59 to 5.78 | 0.822 |

OPEG: Ophthalmology and/or optometry private equity-backed group

**Figure 2.**
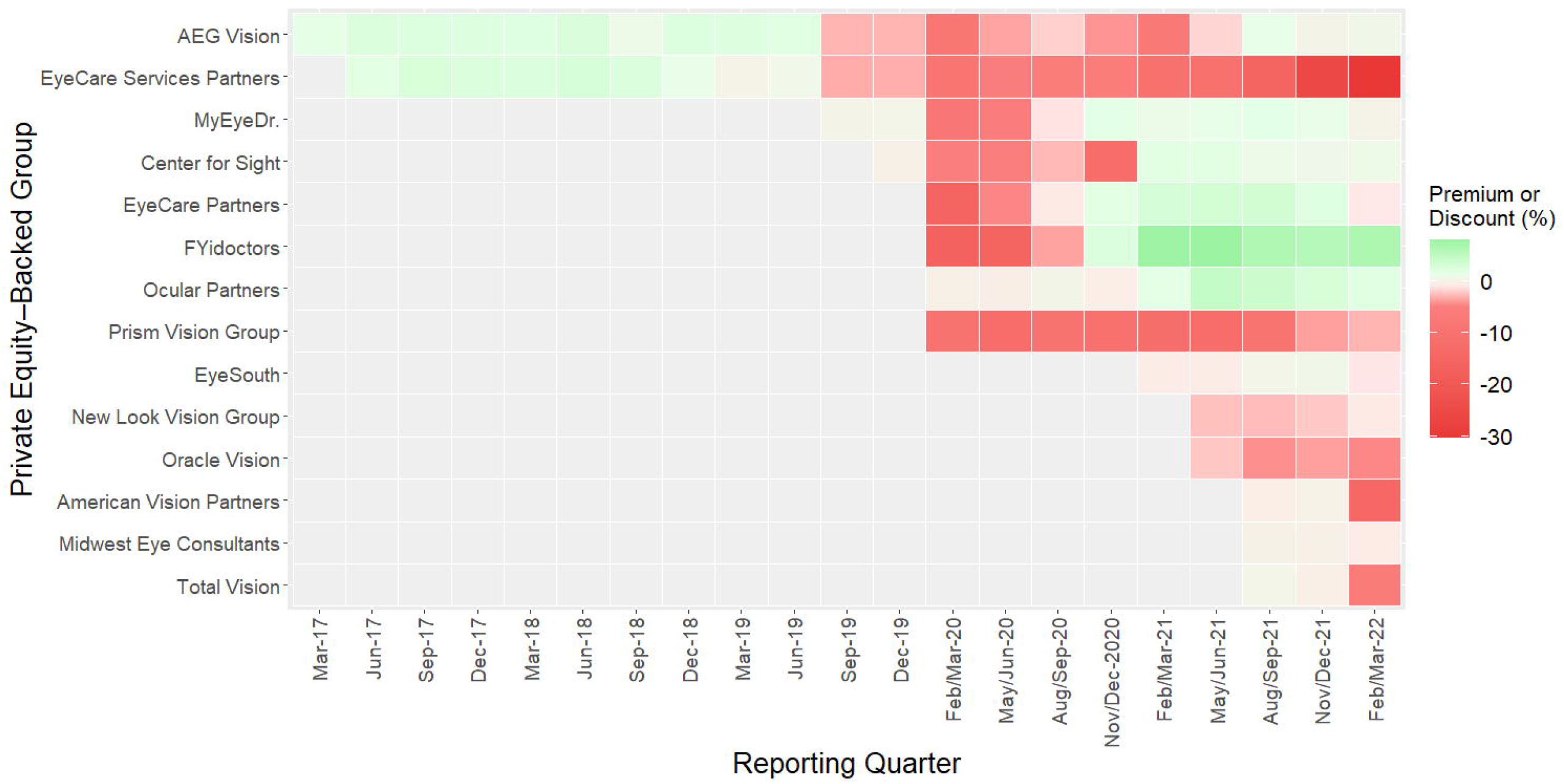
Premium/Discount of PE-Backed Ophthalmology and Optometry Debt Instruments

In addition to this baseline decline in debt valuations, the economic environment of the COVID-19 pre-vaccine period (March 2020 – December 2020) contributed to an excess (additional) 4.93% decrease in valuations (95% CI: −8.63 to −1.24, P = 0.010) when compared to pre-pandemic (March 2017 – December 2019). During this pandemic pre-vaccine period, 100% of OPEGs reported an average discounted debt valuation (Table 2). Three OPEGs, EyeCare Partners, MyEyeDr., and FYidoctors were able to return to premium debt valuations by November/December of 2020. This economic shock on debt trends stabilized by the post-pandemic period (February 2021 – March 2022), with no significant change in excess valuations when compared to pre-pandemic levels (0.60, 95% CI: −4.59 to 5.78, P = 0.822). Despite stabilization of excess debt valuations, the baseline decline in valuations persisted over time. During the pandemic-post vaccine period, 1213 practices (40.5% of all practices) associated with 9 OPEGs reported an average discounted debt valuation. This represents a 25-fold increase in discounted practices when compared to the pre-pandemic period.

Acquisition of debt instruments increased over time (increasing debt assumed by practices), with a focus on debt associated with larger OPEGs with practice locations in multiple states. Compounding the issue of increasing debt, 759 practices associated with 6 OPEGs acquired in the post-vaccine period reported average discounted debt valuations, signaling poor financial performance of these groups following PE investment. Additionally, debt associated with EyeCare Services Partners has plunged into the deep discount classification for the first time (> 20% discount) in the post-vaccine period, indicating extremely high risk of defaulting on debt and potential for bankruptcy.^20^

## Discussion

To our knowledge, our study is the first to assess the debt valuations of PE-backed ophthalmology and optometry practices. Debt associated with the majority of individual OPEGs in 2021 is declining over time, despite a normalization of economic impacts on OPEG debt from pre-pandemic to the pandemic post-vaccine period. Similar findings were appreciated in a study of debt valuations of PE-backed dermatology groups.^9^ Discounted valuations of debt raise concerns about the financial viability of many PE-backed practices, as these valuations indicate practices are less valuable.

Given the significant costs of capital-intensive equipment (e.g. optical coherence tomography, ultra-wide fundus photography, slit-lamps) in combination with declining reimbursements, accepting PE investment is sometimes viewed as a low-risk financial decision for physician owners as. However, PE firms invest little of their own capital and are not vulnerable to major losses if an OPEG declares bankruptcy. In contrast, the OPEG itself is directly responsible for servicing the debt incurred by the PE firm. Holding large debt loads, particularly during periods of economic contraction, can place additional financial stress on OPEGs to service interest payments and can put practices at risk for cost-cutting, restructuring, or even bankruptcy.^5^ The inherently distorted incentive structure in debt acquisition and payoff between practices and a PE firm often creates conflicts in practice management strategy. Factors that have been cited include loss of autonomy, decreased influence on practice culture, increased scope of midlevel practitioners, reduced yearly compensation, and elimination of the right to due process during termination of employment.^21, 22^

In the most extreme situations, a failure to repay debt results in restructuring the terms of the debt agreement or practice bankruptcy. Physicians usually receive a combination of an initial cash payout and equity when selling their practices to PE firms. In restructuring or bankruptcy, debt investors have preference over equity holders in claims on company’s assets.^23^ Thus, physicians typically have the last claim on assets if a company liquidates or recapitalizes to pay off debt, and consequently assume a higher level of risk than debt investors when their practice is acquired. Unfortunately, early career physicians shoulder the highest level of financial risk. After a practice is acquired by an OPEG, they typically do not receive the initial cash payoff that practice partners receive and they work at a reduced income potential.^24^ An increasing number of early career physicians are entering the field as autonomy and ownership continue to diminish, which may further impact physician leverage in future PE negotiations.^25^

Skeptics of the PE model further point to examples in which short term cash flows have been prioritized at a detriment to not only physician autonomy, but also patient outcomes. Physicians are provided financial incentives by the PE firm that may lead to over or under-treatment of patients; PE-backed dentistry groups were found to bill for an increased volume of procedures regardless of medical necessity.^26^ PE-backed nursing homes exhibit an increased short-term mortality rates which may be related to lowered nursing-staff-to-resident ratios and the diversion of patient care funding, both strategies used by PE firms to increase revenue margins.^27^ PE investment in healthcare has even entered the public arena, with articles in the New Yorker as well as in Bloomberg Businessweek based on concerns surrounding patient outcomes and medical decision making in addition to potential anti-trust issues.^28-30^

Initial findings in ophthalmology have been inconclusive, with some articles indicating minimal changes to patient care and others indicating a rising movement toward short-term profits, such as prescribing more expensive medications.^31^ Recently, a difference-in-differences study of PE–acquired dermatology, gastroenterology, and ophthalmology physician practices and independent practices found the former was associated with differential increases in allowed amount and charges per claim, volume of encounters, and new patients seen, as well as some changes in billing and coding.^32^ Further data is required to evaluate PE-backed investments in ophthalmology and optometry, such that quality of care does not decline.

Despite falling valuations and skepticism, PE involvement in eye care is growing at an exponential rate. Investment activity began in the early 2010s, and the market share of PE in optometry & ophthalmology now stands at 16.5% and 14.5%, respectively.^33^ Debt instruments held in BDC portfolios from December 2020 to March of 2022 represent a 167% increase from the 12 OPEG debt instruments reported pre-pandemic. The majority of the private equity deals occurring in ophthalmology and optometry are still primary transactions – a practice or group of practices being sold directly to a PE firm. Secondary transactions, when a PE firm sells all or part of its ownership interest, started in 2018 when EyeCare Services Partners was sold for an undisclosed amount.^34^ The next secondary transaction occurred in 2019, when EyeCare Partners (the largest eye care group in the U.S.) was sold for 2.2 Billion USD by FFL Partners.^35^ During the 4-year hold period, FFL Partners produced a 65% annualized return, outperforming the 4.9% compound annual growth rate for outpatient services between 2016 and 2019 over 13.3 times.^35, 36^ Additionally, in September 2022, Olympus Partners agreed to acquire EyeSouth Partners from Shore Capital in a transaction valued at close to $1 billion.^37^ Strategies such as consolidation, used to increase practice valuations for secondary transactions, indicate a short-term focus on practice growth followed by recapitalization in contrast to physicians and patients, who have a longer term relationship with each other.

There are several limitations of this analysis. First, debt held by BDCs only represents a portion of loans used in the acquisition of OPEGs; additional transparency is required to capture the true magnitude of private debt held by all OPEGs. Second, publicly reported fair value of debt is tabulated by individual BDC firms and reported to the SEC; inaccuracies may exist in these calculations. Third, debt included in this study only includes companies reporting debt in 2021, and the results may not be able to be extrapolated to debt held and repaid by companies in previous years. Lastly, BDCs holding debt instruments for non-PE-backed optometry & ophthalmology corporations were excluded from the study, potentially underestimating debt devaluations in the specialty at large.

Private equity in ophthalmology is a relatively new field of investment, consequently, there is currently little evidence surrounding ramifications of PE ownership on the longer-term financial outlook or patient outcomes of acquired practices. This study’s findings of decreased practice valuations with increasing debt, along with the growing rise of PE in both the literature, financial news, and popular press, raise significant concerns on the impact PE is having on ophthalmology and medicine as a whole, physicians, practices, and most importantly, patients. Future studies should examine the impact of the high inflation and rising interest rates on debt valuations of OPEGs as private markets tend to lag public markets. Additionally, studies should focus on trends in secondary transactions of OPEGs on the financial health of these groups, as well as providing quantitative evidence on the effects of PE investments on physician autonomy, outpatient procedure volumes and patient outcomes.

## Data Availability

All data produced in the present study are available upon reasonable request to the authors

